# Anatomical and spectral foundation of skin pigmentation bias in optical oximetry

**DOI:** 10.64898/2026.11.22.26368599

**Authors:** Felix Wachter, Moritz Schillinger, Adrian Buehler, Benjamin Stoecklein, Hannah Vogt-Wolz, Jan Nabo, Charlene Reichl, Neal Burton, Henriette Grieshaber-Bouyer Mandelbaum, Joachim Woelfle, Heiko Reutter, Adrian P. Regensburger, Angelos Karlas, Ulrich Rother, Katharina Breininger, Ferdinand Knieling

## Abstract

**IMPORTANCE:** Monitoring hemoglobin oxygen saturation is a cornerstone of patient care across a wide range of clinical settings, with pulse oximetry remaining one of the most widely used technologies. Recent studies have identified skin pigmentation–related inaccuracies in these measurements; however, the biological and optical mechanisms underlying these limitations remain incompletely understood.

**OBJECTIVE:** To investigate the tissue compartments and wavelengths that drive skin pigmentation– related bias in oximetry using depth-resolved optoacoustic (OA) imaging.

**DESIGN, SETTING, AND PARTICIPANTS:** Prospective single center observational imaging study of n=46 healthy volunteers representing all 6 Fitzpatrick skin types (FST).

**EXPOSURES:** OA imaging of the tibialis anterior region during physiological oxygenation challenges induced by vascular occlusion, reperfusion, and exercise.

**MAIN OUTCOMES AND MEASURES:** A total of 3016 spatially resolved scans were obtained. Tissue compartments were segmented using a pretrained neural network. OA spectra, oxygen saturation estimates, responses to physiological interventions, and associations with skin pigmentation were evaluated across 41 wavelengths from 700 to 1100 nm.

**RESULTS:** Skin OA spectra closely mirror the optical absorption characteristics of melanin, and OA signal intensities are strongly associated with FST, demonstrating a substantial influence of pigmentation on superficial oximetry measurements. Epidermal oxygen saturation estimates exhibit no reliable changes during physiological interventions, especially in darkly pigmented skin. Skin signals show pronounced associations with melanin-sensitive wavelengths, enabling FST classification based on optoacoustic skin signals. Pigmentation-related bias is greatest at shorter wavelengths and progressively decreased at longer wavelengths. In contrast, oxygenation measurements obtained from skeletal muscle are less affected by skin type, although absolute values remain higher in dark skin, and show the strongest responses to vascular occlusion, reperfusion, and exercise. Depth-resolved OA imaging demonstrates that physiologically meaningful oxygenation information emerges only after propagation beyond the avascular epidermis into deeper vascularized tissue compartments.

**CONCLUSIONS AND RELEVANCE:** A better understanding of how skin pigmentation affects light-tissue interactions will enable appropriate anatomical targeting, optimized wavelength selection, and correction for optical attenuation. Together, these approaches may offer a promising strategy for mitigating skin-pigmentation-related bias in oximetry.

**Key Points:** *Question:* What are the anatomical and spectral determinants of skin pigmentation–related bias in oximetry?

*Findings:* In this prospective observational study of 46 healthy volunteers across all 6 Fitzpatrick skin types, optoacoustic imaging enabled spatially resolved oxygenation mapping and assessment of tissue-specific absorption across 41 wavelengths. Skin pigmentation was strongly associated with superficial optoacoustic signal intensities and oxygenation measurements, with the greatest bias at shorter wavelengths. In contrast, skeletal muscle oxygenation measurements were less affected by skin pigmentation and showed robust responses to physiological oxygenation challenges.

*Meaning:* Anatomical targeting of deeper vascularized tissue compartments and optimized wavelength selection may help mitigate skin pigmentation–related bias in oximetry.

## INTRODUCTION

Tissues rely on continuous oxygen supply for sufficient function and survival. In clinical practice, oximetry – the measurement of oxygen content or saturation in biological tissue – is a cornerstone technology in modern patient monitoring. It is recognized as an essential component of safe surgical care and is explicitly included in the WHO Surgical Safety Checklist^1^, and its use extends throughout everyday clinical practice, including emergency medicine, critical care, inpatient ward monitoring, and ambulatory care.

Oximetry technologies are typically based on optical imaging and sensing that assess either systemic oxygenation, as in pulse oximetry, or regional tissue oxygenation, as in near-infrared spectroscopy (NIRS). There is growing evidence that these light-based technologies may be subject to important technological and biological limitations, including performance disparities associated with skin pigmentation – often referred to as “racial bias”.^2–5^ In pulse oximetry, hypoxemia is more likely to be undetected in patients with darker skin pigmentation^2^, and this measurement error has been associated with adverse clinical outcomes, including increased mortality.^3^ Low perfusion is another important factor affecting device performance, particularly in acutely ill patients, in whom accurate measurements are even more critical for timely clinical decision-making^6^.

In response to these concerns, the FDA has issued draft guidance with strengthened recommendations for the clinical validation of pulse oximeters, emphasizing substantially larger study populations, standardized characterization of skin pigmentation, and demonstration of comparable performance across diverse skin tones.^7^ While these measures may improve the detection and quantification of performance disparities, they do not directly address the underlying causes of measurement bias. Progress toward overcoming these technological and biological limitations has been constrained by an incomplete understanding of the optical and physiological mechanisms underlying oximetry measurements. In particular, the tissue compartments contributing to the detected signals, the influence of skin pigmentation on light propagation and signal formation, and the wavelength dependence of these effects remain incompletely characterized, especially in humans. A deeper mechanistic understanding of these factors is required for the development of more accurate, robust, and equitable oximetry technologies.

Unlike conventional oximetry techniques, which typically provide an integrated measure of oxygen saturation across bulk tissue, optoacoustic (OA) technologies provide depth-resolved information on optical tissue characteristics.^8–12^ This capability allows oxygenation signals to be localized and assigned to specific tissue compartments^13,14^ and may provide a unique opportunity to investigate the biological and physical factors underlying measurement bias. Using OA imaging, we present spatially resolved oximetry in human subjects across the full spectrum of skin pigmentation. We identified and quantified wavelength-dependent pigmentation effects on optoacoustic signals and oxygen saturation estimates and determined the tissue compartments contributing to these effects. Our findings provide insight into the origins of oximetry bias and establish a foundation for the development of the next generation of oximetry technologies.

## METHODS

### Study Population

The study was conducted in accordance with the ethical principles of the Declaration of Helsinki and was approved by the Ethics Committee of the Friedrich-Alexander-Universität Erlangen-Nürnberg (approval number 24-167_1-B). All participants provided written informed consent before enrollment. The study was registered at ClinicalTrials.gov (ID NCT06541496). Participants were prospectively recruited for this clinical study through public notices displayed throughout the city, with particular emphasis on achieving representation across the full range of Fitzpatrick skin types (FST I–VI). Eligible individuals were adults aged 18 years or older who were able to provide written informed consent and comply with all study procedures. FST was determined using a reference color scale by the examiner. Exclusion criteria included pregnancy or breastfeeding; active skin disease, infection, wounds, burns, tattoos, or significant scarring at the imaging site; or any medical condition that, in the opinion of the investigators, could interfere with study participation or the interpretation of imaging findings. Participants were also excluded if they were unable to remain still during image acquisition or if technical factors precluded acquisition of images of sufficient quality for analysis.

### Data Collection

Baseline demographic and clinical characteristics were recorded for all participants, including age, sex, body weight, body mass index (BMI), FST, relevant medical history, concomitant medications, and pre-existing medical conditions.

### Imaging Protocol and Data Processing

Participants underwent standardized OA imaging at the tibialis anterior muscle. Imaging was performed at five time points: resting baseline, during transient tissue oxygen desaturation induced by inflation of a pneumatic cuff, during reperfusion (hyperperfusion) induced by the release of the cuff, during return to baseline, and following standardized physical exertion. Multiple image acquisitions were obtained during each phase. All imaging was performed using the CE-certified Acuity MSOT system (iThera Medical GmbH, Munich, Germany). For each acquisition, OA data were collected at 41 discrete wavelengths ranging from 700 to 1100 nm in 10-nm increments. Co-registered B-mode ultrasound images were acquired simultaneously for anatomical reference and subsequent tissue segmentation.

Raw OA data were reconstructed using a back-projection algorithm^15^ and a tissue segmentation mask was generated on the co-registered ultrasound images using a convolutional neural network trained on an independently annotated dataset^16^. The model classified each image pixel into one of four anatomical tissue compartments: skin, subcutaneous adipose tissue, fascia, or skeletal muscle. Chromophore signals were calculated by linear spectral unmixing to generate spatially resolved distributions of oxygenated hemoglobin (HbO₂) and deoxygenated hemoglobin (Hb). Oxygen saturation (SO₂) was calculated on a pixel-wise basis according to:

SO_2_ = HbO_2_ / (Hb + HbO_2_).

Pixels yielding negative Hb or HbO₂ values were considered physiologically implausible and were excluded from further analysis. For post-processing, images were clipped to a depth of 120 pixels and a centrally positioned lateral region spanning 200 pixels. Single wavelength signal and unmixed hemoglobin signal were quantified for a region of interest within each tissue segment (10 pixels in depth and 10 pixels in width for the skin, 20 pixels in depth and 50 pixels in width for the muscle). To generate oxygenation masks combining multiple participants, tissue boundaries were extracted from the segmentation masks and converted into column-wise thickness profiles. These profiles were aggregated across participants to generate a common anatomical template representing the FST cohort. To facilitate spatially resolved group analyses, individual SO₂ maps were transformed from subject-specific anatomical geometries into the common template using boundary-constrained layer-wise resampling. This normalization approach enabled direct comparison of homologous tissue regions across participants despite interindividual differences in tissue thickness and morphology. Subject-level oxygenation change maps (ΔSO₂) were calculated by subtracting the median baseline SO₂ map from the median occlusion-phase SO₂ map, so that negative values indicate desaturation. These maps quantified the spatial extent and magnitude of oxygen desaturation induced by vascular occlusion. Group-level delta oxygenation maps were generated by computing the pixel-wise median of all subject-level delta maps.

### Statistical Analysis

For analysis, participants were grouped as light (FST I–II), medium (FST III–IV), or dark skin (FST V–VI). Demographic, clinical, and imaging variables were summarized using descriptive statistics. Continuous variables are reported as mean and standard deviation (SD) or median and interquartile range (IQR), as appropriate, and categorical variables as counts and percentages. Parameters were compared across FST groups using nonparametric statistical tests. Changes in oxygenation over different time points within individual participants were assessed using the Friedman test for repeated measures, followed by multiple-comparisons testing for post hoc pairwise comparisons. Associations between FST and imaging parameters were assessed using Spearman’s rank correlation. Receiver operating characteristic (ROC) curve analyses were performed to evaluate the ability of imaging parameters to discriminate between individual Fitzpatrick skin types (FST I– VI) using pairwise comparisons and between FST I–III and FST IV–VI, with corresponding areas under the curve (AUCs) calculated for each comparison. All statistical tests were two-sided, and a significance threshold of P < .05 was considered statistically significant. Analyses were performed using GraphPad Prism (version 10, GraphPad Software, Boston, MA, USA).

## RESULTS

### Study Population

Fifty participants were enrolled and underwent the imaging protocol. After image acquisition and quality-control procedures, 4 participants were excluded from the final analysis because of technical failures causing incomplete imaging data (n=3) or a confounding factor (high blood pressure, n=1). The final study cohort consisted of 46 participants representing all Fitzpatrick skin phototypes (**Figure 1**) and a total of n=3016 scans were obtained. Seven participants (15.2%) were classified as FST I–II, 25 (54.3%) as FST III–IV, and 14 (30.4%) as FST V–VI. Baseline demographic characteristics across skin-type groups are displayed in Table 1. Peripheral oxygen saturation measured by pulse oximetry was similar across groups (**Table 1**).

**Figure 1.**
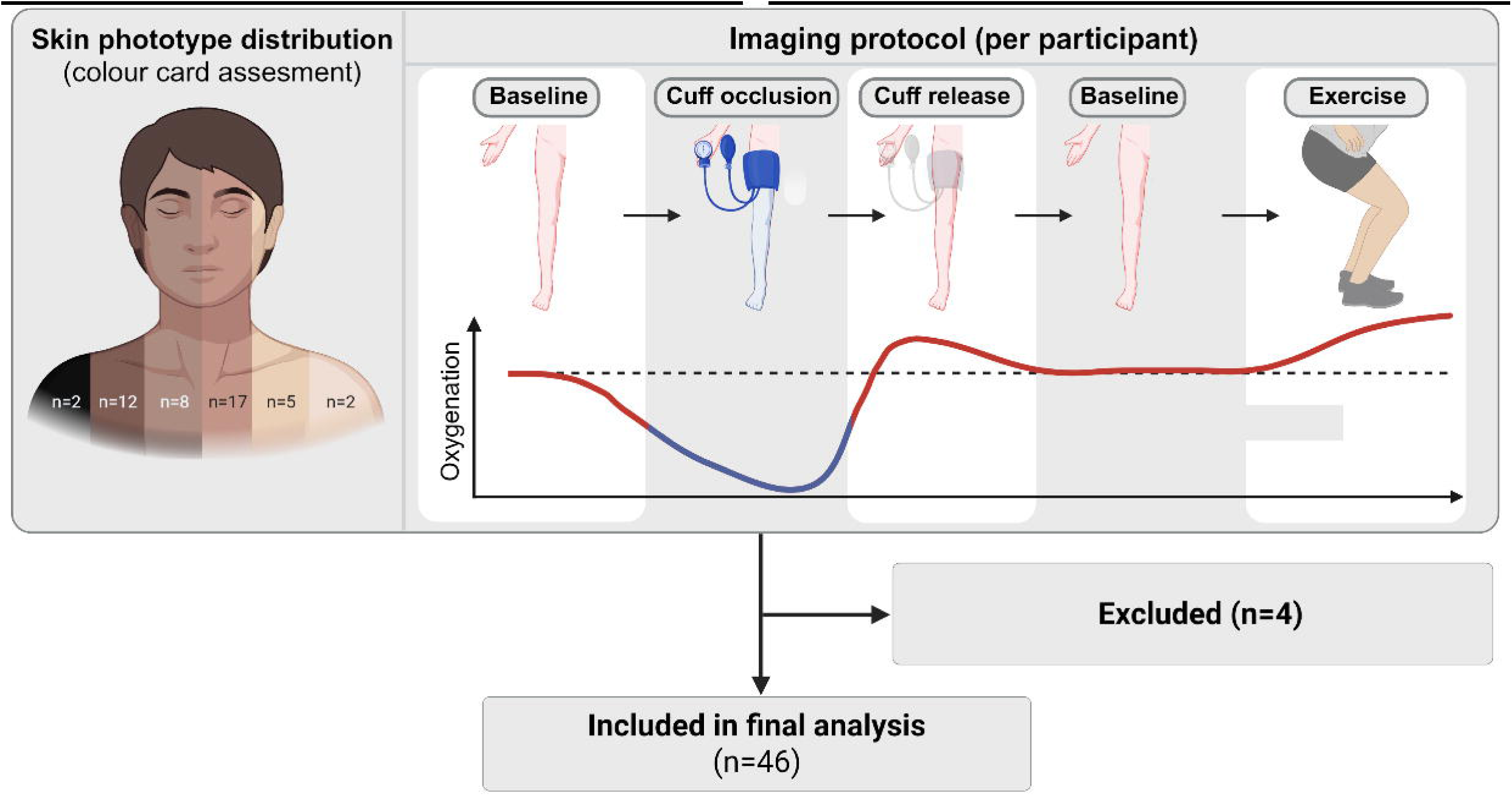
Imaging Protocol. Flow diagram of imaging protocol and analysis. All participants underwent OA imaging of the tibialis anterior muscle during baseline conditions, transient vascular occlusion induced by pneumatic cuff inflation, reperfusion after cuff release, return to baseline, and standardized exercise.

**Table 1.** Participant baseline demographics.

|  | **Fitzpatrick I–II (n = 7)** | **Fitzpatrick III–IV (n = 25)** | **Fitzpatrick V–VI (n = 14)** | **Overall (n = 46)** |
|---|---|---|---|---|
| **Age, mean (SD), y** | 26.6 (6.7) | 29.3 (6.9) | 29.4 (5.6) | 28.9 (6.6) |
| **Female sex, No. (%)** | 6 (85.7%) | 17 (68.0%) | 6 (42.9%) | 29 (63.0%) |
| **Weight, mean (SD), kg** | 59.6 (12.7) | 71.3 (15.8) | 74.0 (12.1) | 70.4 (15.0) |
| **BMI, mean (SD), kg/m²** | 21.3 (4.9) | 24.9 (4.6) | 26.0 (3.7) | 24.7 (4.6) |
| **Pulse oximetry, mean (SD), %** | 97.9 (1.2) | 98.1 (1.3) | 97.8 (0.9) | 98.0 (1.2) |

### Safety

All procedures were well tolerated and none was discontinued. No skin irritation, erythema, tissue injury, or other local adverse effects were observed at the measurement site.

### Anatomical Feasibility of Spatially Resolved Oximetry

To simulate different oxygenation conditions, five time points were examined, encompassing both low perfusion and hyperperfusion states (**Figure 1**). By calculating the differences (delta SO₂) between time points, spatially resolved oxygenation maps were generated to identify anatomical regions suitable for quantitative oximetry (**Figure 2A**). Epidermal signals depended strongly on skin pigmentation, whereas deeper compartments, particularly muscle tissue, still showed a measurable response (**Figure 2A, B**). Changes in oxygenation could not be measured in the skin layer (P > .05 for all time points, **Figure 4**). This was the case for participants of all three skin type groups. However, a small band of suitable targets for oximetry was identified underneath the epidermis in the dermal layer (**Figure 2A**). In contrast, deeper muscle tissue still retained a detectable hemoglobin spectrum (**Figure 3B**) and even slight changes in oxygenation could be detected, including in participants with darker skin color **(Figure 4**).

**Figure 2.**
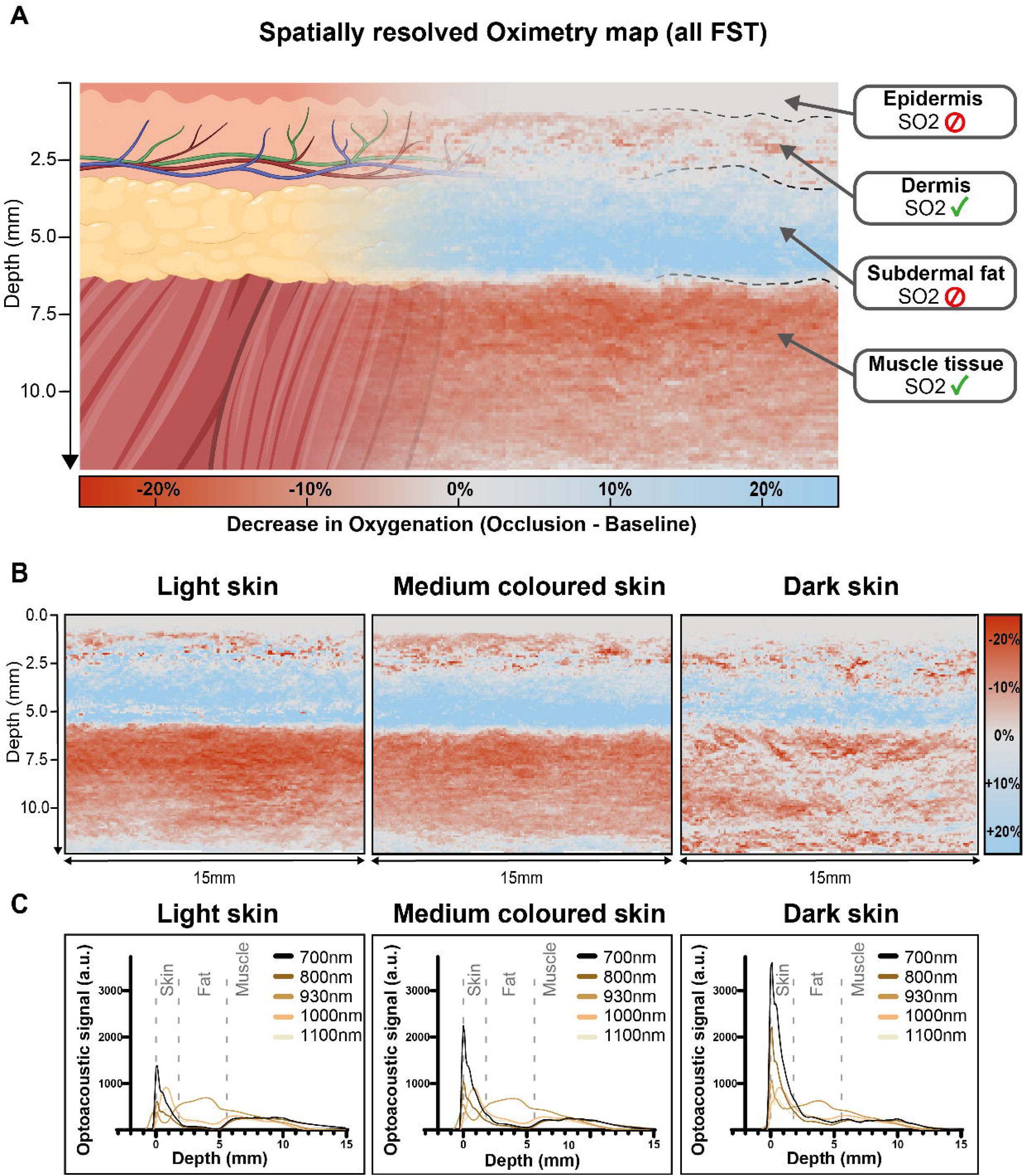
Spatially resolved maps to illustrate where oximetry is feasible. The maps show the median change in oxygenation during leg occlusion compared to baseline across all participants (A) and in subgroups stratified by skin color (B). Red indicates a decrease in oxygenation and therefore identifies regions that are particularly suitable for oximetry. In contrast, white and blue regions (epidermis and subdermal fat layer, respectively) indicate areas in which a decrease in oxygenation could not be reliably measured. C) Depth profiles of the optoacoustic signal at selected wavelengths (700–1100 nm) in light, medium-colored and dark skin. The superficial skin signal increases with skin color and is highest at short wavelengths, whereas signals in deeper tissue are comparable across groups.

**Figure 3.**
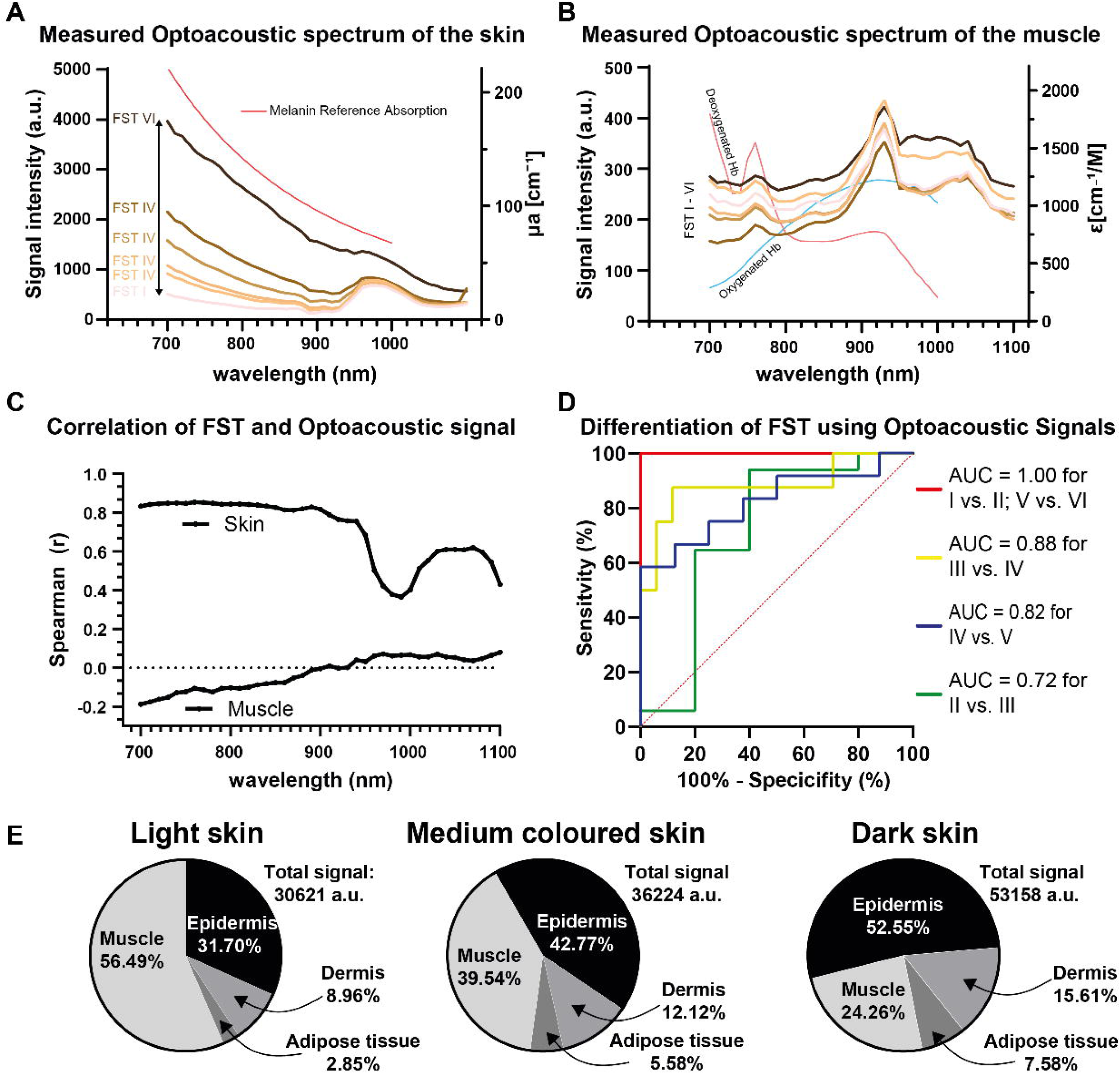
Melanin effect on the optical spectrum. A) The signal across the 41 measured wavelengths closely resembles the absorption spectrum of melanin (red line: literature reference^31^). B) Absorption spectra measured in muscle across the different skin-color groups. C) Correlation between wavelength-specific signals measured in the skin and muscle and skin color. D) Diagnostic discriminability between skin-color groups based on the 760 nm optoacoustic signal measured in the skin. E) Origin of the measured signal. Although the epidermal layer is less than one millimeter thick and does not pose a suitable target for oximetry, most of the signal stems from this tissue in dark-skinned participants. Shown is the bulk signal (Total signal) and the corresponding tissue types across skin colors.

**Figure 4.**
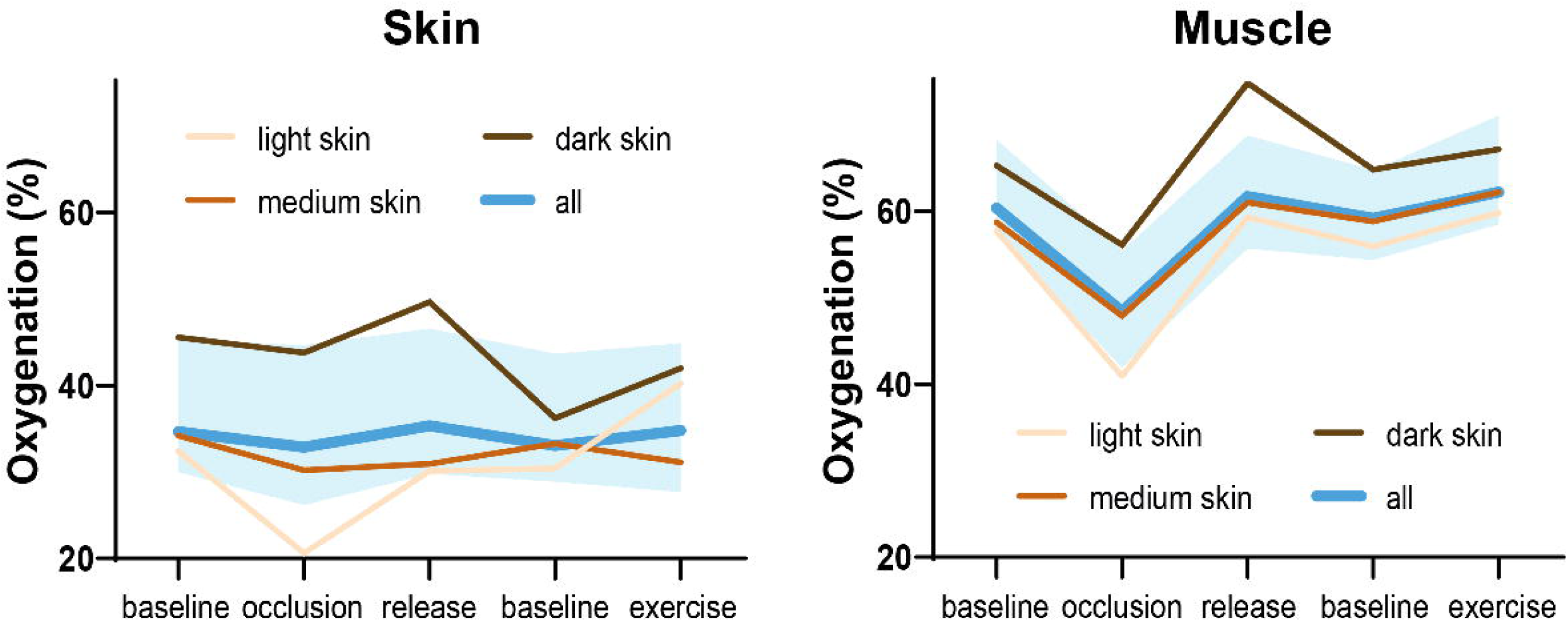
Oxygenation measurements in the skin and in the muscle. The overestimation of oxygenation in participants of darker skin color is evident in both the skin and muscle tissue. However, while changes in oxygenation induced by occlusion, reperfusion following release, and exercise are clearly detectable in the muscle, they are not reliably detectable in the skin layer. The blue background represents the interquartile range (IQR) of the blue line, which indicates the median oxygenation across all participants.

### Melanin Dominates the Optoacoustic Signal of the Skin

Spectral analyses revealed layer-specific optical responses across the investigated wavelength range, with shorter wavelengths exhibiting stronger attenuation.

The spectral analysis of the superficial skin layer showed that the measured signal closely resembled the known absorption spectrum of melanin, signal intensities matched FST and lacked detectable features of Hb and HbO₂ (**Figure 3A**). Signal intensity in the skin demonstrated a strong positive correlation with FST at shorter wavelengths (r = 0.8 below 920 nm) (**Figure 3C**). At 700 nm, signal intensity increased stepwise with skin type (8-fold increase from FST I to FST VI), and the differences between groups narrowed towards longer wavelengths. Depth profiles showed the same behavior: the superficial skin peak was highest at 700 nm and increased from light (approximately 1,400 a.u.) over medium-colored (approximately 2,200 a.u.) to dark skin (approximately 3,600 a.u.), whereas signals beyond the skin–fat boundary were of comparable magnitude across groups (**Figure 2C**). In contrast, the muscle spectrum displayed the characteristic hemoglobin features with a peak at approximately 930 nm, retained its spectral shape in all skin-color groups and differed only by a comparatively small offset (**Figure 3B**).

Skin-derived melanin signals discriminated FST I–III from FST IV–VI with high accuracy (AUC, 0.953; P < .0001). Pairwise comparisons yielded AUCs of 1.00 for FST I vs. II and V vs. VI, 0.88 for III vs. IV, 0.82 for IV vs. V and 0.72 for II vs. III (**Figure 3D**). In muscle, the correlation of signal intensity and FST was weak and slightly negative at short wavelengths (r −0.1 to −0.2) and approached zero above 900 nm (**Figure 3C**).

### Contribution of Tissue Compartments to the Total Signal

The fraction of the bulk signal (total signal) originating from the epidermis increased from 31.7% in light skin to 42.8% in medium-colored and 52.6% in dark skin, whereas the fraction originating from skeletal muscle decreased from 56.5% to 39.5% and 24.3%, respectively (**Figure 3E**). Dermal (9.0%, 12.1%, 15.6%) and adipose tissue fractions (2.9%, 5.6%, 7.6%) also increased with skin color. The total signal rose moderately by 18.3% in light to medium-colored and by 73.5% when comparing light to dark skin. Calculated from these percentages, the absolute epidermal signal increased approximately threefold between light and dark skin (about 9,700 to 27,900 a.u.), while the absolute muscle signal decreased by roughly a quarter (about 17,300 to 12,900 a.u.). Consequently, in dark skin the avascular, melanin-rich epidermis contributed about twice as much signal as skeletal muscle.

### Oxygenation Changes During Physiological Challenges

Spatially resolved delta maps showed a pronounced decrease in estimated SO₂ in skeletal muscle during cuff occlusion (up to approximately −20%). In contrast, the epidermis showed no reliable change, the dermis only a narrow band of decrease, and the subdermal fat layer an apparent increase of up to approximately +20% (**Figure 2A**). This pattern was reproduced in light and medium-colored skin, whereas in dark skin the muscle response was weaker and spatially less homogeneous (**Figure 2B**). The apparent oxygenation increase in the fat layer can be attributed to a fluence (spectral coloring) artifact, as shown by Monte Carlo simulation^17^ (Supplementary Material).

In muscle, median SO₂ of all participants decreased from approximately 60% at baseline to approximately 48% during occlusion, increased to approximately 62% after cuff release, returned to approximately 59% at the second baseline and reached approximately 62% after exercise (**Figure 4**, right). In contrast to the skin measurements, in the muscle measured oxygen changes compared to baseline were significant during occlusion, reperfusion and exercise (all P < .05). The occlusion-induced decrease was detectable in all skin-color groups (approximately 17, 11 and 9 percentage points in light, medium-colored and dark skin, respectively). However, absolute values in dark skin were shifted upwards by roughly 5–15 percentage points throughout the protocol (approximately 65% at baseline and 75% after release), consistent with an overestimation of oxygenation. In the skin, median SO₂ of all participants remained essentially flat at approximately 33–36% across all five time points, with changes remaining within the interquartile range (**Figure 4**, left). Light skin showed larger but inconsistent fluctuations (approximately 20% during occlusion and 40% after exercise), and dark skin showed consistently higher values (approximately 36–50%) without a clear physiological response pattern.

### Wavelength Dependence of Oxygenation-Related Signal Changes

Depth- and wavelength-resolved analysis of the signal difference between baseline and occlusion showed that, within muscle (below 5.7 mm), the signal decreased during occlusion at wavelengths above approximately 890 nm, most strongly around 920–940 nm and between 1,020 and 1,100 nm. This is in line with the higher absorption of HbO₂ above the isosbestic point at 805 n. Within the fat layer (1.8–5.7 mm), the changes showed the opposite pattern, with a signal decrease at 700–780 nm and an increase at 910–1,100 nm, corresponding to the apparent oxygenation increase described above. Together with the wavelength dependence of the skin signal (**Figure 3C**), these findings indicate that pigmentation-related bias was greatest at short wavelengths and that longer wavelengths in combination with deep, vascularized targets such as skeletal muscle are best suited for oximetry across skin types.

## DISCUSSION

Recent evidence demonstrating systematic inaccuracies of pulse oximetry in individuals with darker skin pigmentation has raised substantial concerns regarding equity in clinical monitoring and patient care. Multiple studies have shown that conventional pulse oximeters may overestimate arterial oxygen saturation in patients with darker skin tones, potentially delaying recognition of hypoxemia and influencing clinical decision-making.^3,18–20^ Although these observations have prompted regulatory scrutiny and renewed interest in equitable monitoring technologies, the biological and technical mechanisms underlying skin-pigmentation bias remain incompletely understood. A major limitation of conventional oximetry technologies like pulse oximetry or NIRS is its lack of spatial resolution, which precludes identification of the tissue compartments contributing to measurement error.

Previous studies identified melanin as an important determinant of optical fluence distribution, thereby confounding quantitative signal measurements.^21,22^ Advances in image reconstruction and signal processing may partially mitigate skin-tone-related degradation of image quality.^23,24^ Using spatially resolved OA oximetry, we provide a mechanistic explanation for inaccuracies in oximetry. We demonstrate that the epidermal compartment is dominated by melanin absorption and lacks a sufficiently measurable hemoglobin target, resulting in oxygenation estimates that were strongly associated with pigmentation but largely insensitive to physiological oxygenation changes. These findings support the view that light propagation beyond the avascular epidermis (0.06–0.6 mm) is required to access vascularized tissue compartments that contain physiologically meaningful oxygenation information.^25,26^ However, we found that in participants with FST V–VI, more than 50% of the bulk OA signal is generated by the epidermal layer.

The first hemoglobin-containing vascular structures within the superficial dermis may themselves represent a challenging target for oxygenation assessment because their oxygenation is strongly influenced by local physiological processes, including thermoregulation, vasomotor tone, and endothelial regulation.^27^ Furthermore, evidence from other optical technologies suggests that clinically relevant pulse oximetry signals originate predominantly from pulsatile arteriolar and arterial compartments located within deeper dermal and subcutaneous tissues rather than from the superficial capillary plexus alone.^28^

Whereas previous studies largely relied on simulations, static observations or phantoms,^29,30^ the present work provides dynamic, compartment-resolved evidence that accurate oxygenation assessment depends on appropriate anatomical targeting. Our findings offer a plausible mechanistic explanation for the systematic overestimation of oxygenation reported in individuals with higher skin pigmentation: superficial melanin-rich tissue can dominate optical signals while contributing little physiologically relevant hemoglobin information. In contrast, deeper vascularized tissue compartments exhibited robust oxygenation responses and reduced susceptibility to pigmentation-related bias.

This study has several limitations. Measurements were performed in healthy volunteers during experimentally induced, moderate desaturation and may not represent clinical conditions such as hypoxemia, low perfusion, or circulatory compromise. The sample size was small, particularly in the light-skinned group; subgroup findings should be confirmed in larger, more balanced cohorts. Sex and body mass index also differed between skin-type groups (Table 1), which may have influenced subcutaneous tissue thickness and light propagation independently of pigmentation. Analyses were restricted to a single anatomical region and tissue thickness and vascularization may differ at common oximetry sites such as the finger or earlobe. Skin pigmentation was classified by a color-card FST only, a coarse measure that may not capture interindividual variation in melanin content and optical tissue properties and was not validated against objective spectrophotometry. In addition, optoacoustic oxygen saturation estimates were not validated against an independent reference measurement. Finally, findings were obtained with a single OA system and cannot be directly transferred to pulse oximetry or NIRS, which might differ in wavelengths, signal origin, and depth resolution. Our results therefore provide a mechanistic explanation rather than a quantification of bias in these devices and require dedicated validation.

Beyond pigmentation-related bias, our observations may also have implications for patients with impaired perfusion. Conventional pulse oximetry relies on adequate peripheral perfusion to isolate the pulsatile arterial component and may become less reliable in conditions with impaired peripheral circulation, including severe vasoconstriction, shock and hypothermia^6^. Because OA imaging does not depend on a pulsatile signal and can resolve deeper tissue, it might be less affected by such conditions.

By identifying tissue compartments that provide physiologically meaningful oxygenation information and characterizing their wavelength-dependent susceptibility to pigmentation effects, our findings suggest a conceptual framework for improving oximetry through anatomically informed target selection, wavelength choice, and correction for optical attenuation. Ultimately, this may help reduce disparities in oxygenation assessment and foster equity in clinical monitoring across diverse patient populations.

## Data Availability

All data produced in the present study are available upon reasonable request to the authors.

## Acknowledgments

We thank all study participants for their contribution to this project.

## Author contributions

FW, FK, NB, and CR contributed to the conceptualization and design of the study, development of the study protocol, and preparation and submission of the ethics application. CK was responsible for data acquisition and study coordination. FW, MS, JN and AB contributed to data analysis and interpretation. The remaining authors contributed through their respective clinical, scientific, and technical expertise, including methodological advice, support with study implementation, image acquisition and assessment, and critical supervision throughout the study. All authors contributed to the interpretation of the findings and the writing and critical revision of the manuscript. All authors read and approved the final manuscript and agree to be accountable for all aspects of the work.

