## Supplementary material for "Anatomical and spectral foundation of skin pigmentation bias in optical oximetry"

**Regarding the non-physiological increase in oxygenation during leg occlusion in the fat layer:**

To test whether the apparent oxygenation increase in the subcutaneous fat layer during occlusion is an artifact, we used our previously developed tissue-aware fluence modeling framework^31^, which builds on the SIMPA toolkit^16^. We applied it to the average layer geometry of our tissue atlas (skin, fat, muscle) to generate a realistic digital twin, and simulated multispectral fluence and initial pressure with Monte Carlo light transport at 700, 800, and 900 nm for baseline and occlusion. Occlusion was modeled by lowering blood oxygenation in the skin (0.8 → 0.2) and muscle (0.6 → 0.3), while the optical properties of fat were kept identical to baseline. Although the true fat oxygenation was unchanged, the simulated delta maps showed a clear signal decrease in the fat layer at 700 nm and an increase at 900 nm, matching the pattern observed in vivo. This signal arises because deoxygenation of the overlying skin increases its absorption at 700 nm (where Hb absorbs more strongly than HbO₂) and decreases it at 900 nm (where HbO₂ dominates). The skin attenuates the two wavelengths differently, which changes the spectrum of the light reaching the fat. Spectral unmixing then misattributes this change in fluence to an increase in fat HbO₂. These results indicate that the physiologically implausible fat-layer changes are fluence (spectral coloring) artifacts and not true local oxygenation changes.

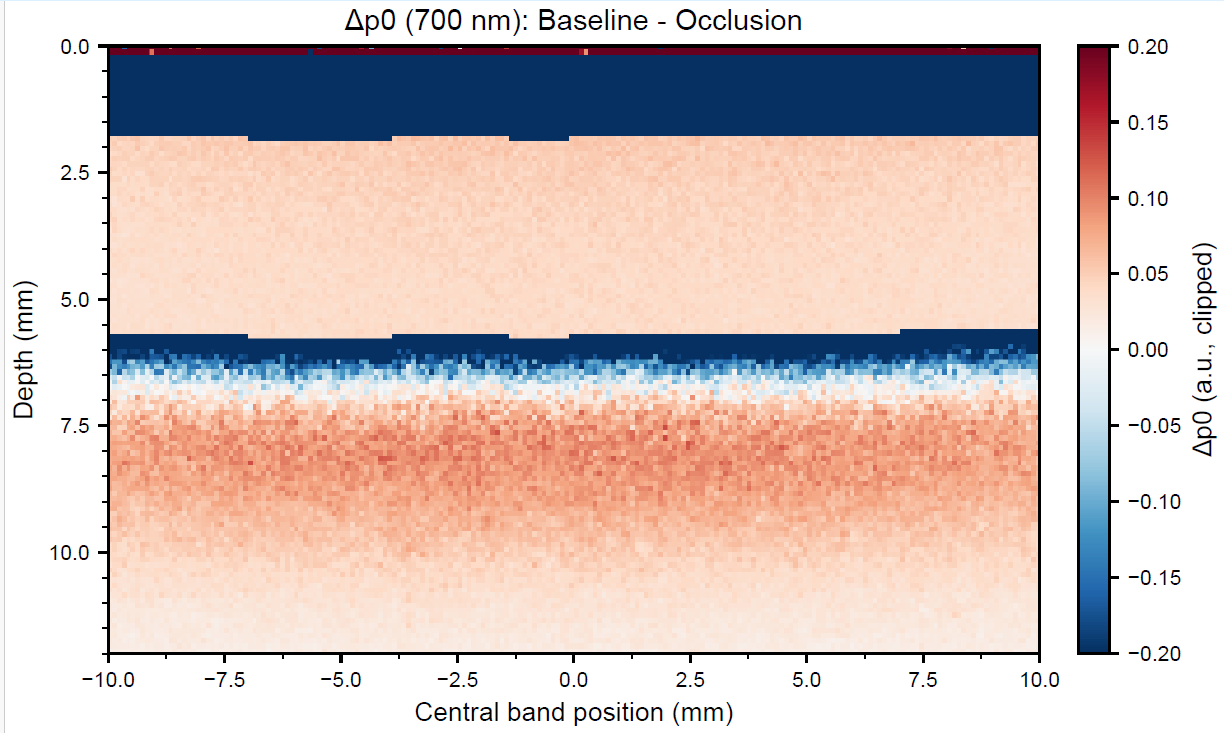

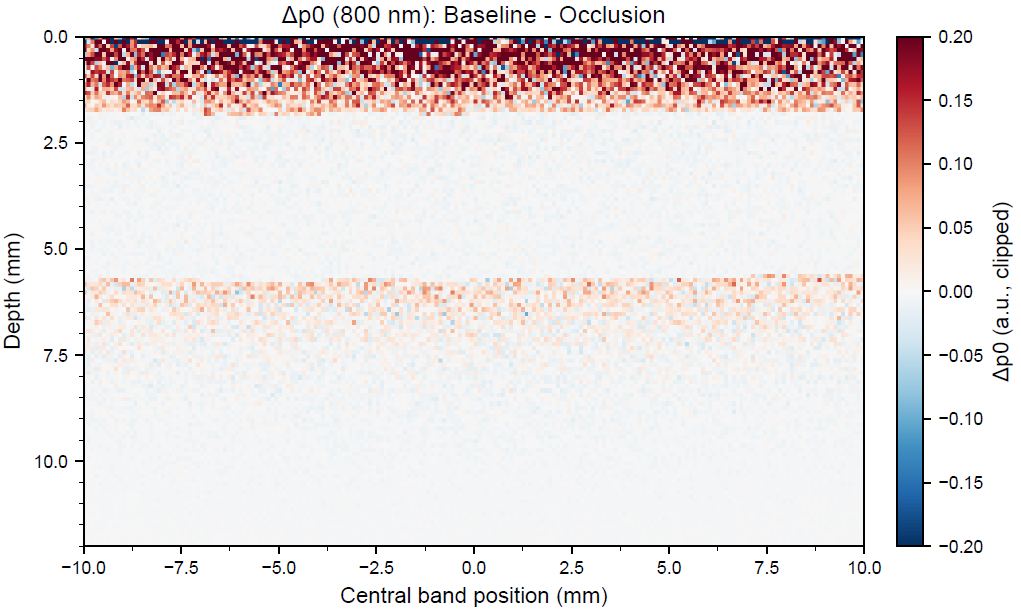

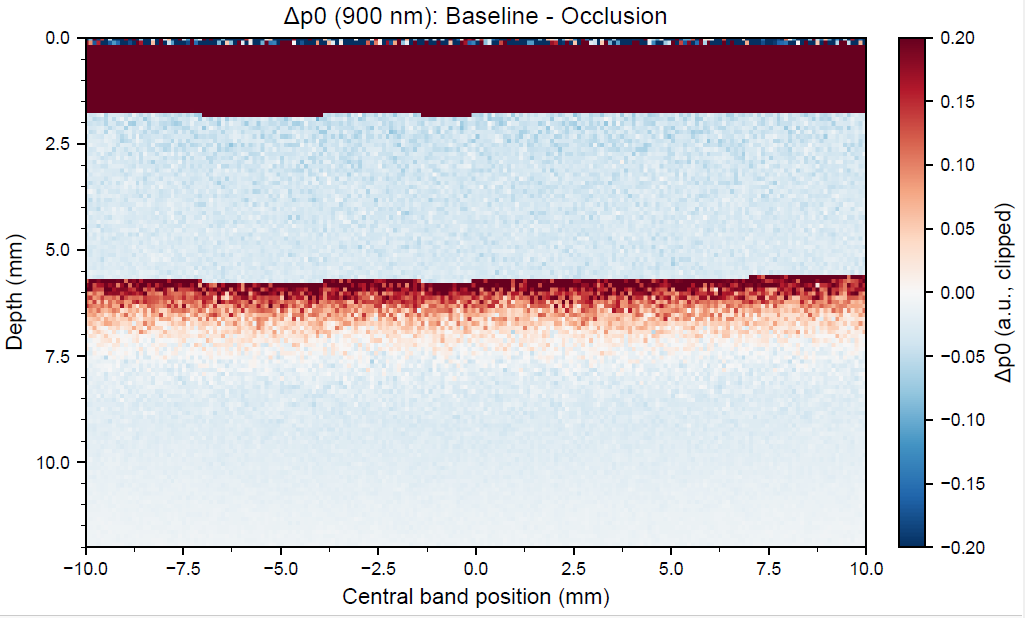
